# Point-of-care thermal imaging tool assisting in the development of imaging biomarker as pre-diagnostics of graves’ disease

**DOI:** 10.1101/2022.07.05.22277045

**Authors:** Vaishali Sharma, Vandana K Dhingra, Ashok Kumar, Snehlata Shakya, Mayank Goswami

## Abstract

This article presents the development of a tool to address the scarcity of clinically validated datasets for Thyroid related abnormalities. Infrared thermography Images are taken from each of the sixty persons of different age groups and gender. Scintigraphy and standard thyroid blood test results are used to categorize these persons into thirty-three females and thirteen males suffering from graves’ disease. Eleven females and three males are found to be in healthy conditions and used as control. An Artificial Intelligence algorithm is used to automatically segment and extract the histogram-associated information within the thyroid and cheek region from the collected images. A very simple and novel imaging biomarker is found to be moderately correlated w.r.t age and gender. A smartphone app integrated with a dedicated smartphone-based compact IR camera add-on is developed and deployed in a clinical environment to enrich the analysis. This point-of-care tool is expected to categorize healthy cases from patients automatically. It is to reduce the ethical burden on clinicians’ shoulders before recommending radioactive contamination-prone Scintigraphy and/or expensive and relatively slower thyroid blood tests. If adopted, such preliminary tests will save costs on patients’ end and burden at pathology labs, especially in densely populated countries such as India.

## Introduction

Early, easy, and cost-effective disease detection is one of the primary goals in healthcare research. Harmful treatment paths with inherent side effects or loss of life (in the worst case) can be avoided if the condition is detected in its initial stages.

Modern-day clinical diagnostics, besides invasive tests, are supplemented by non-invasive imaging modalities that have their own specific and limited field of applications [1]–[4]. Magnetic resonance imaging (MRI), X-ray radiography, scintigraphy, X-Ray computerized tomography (CT), positron emission tomography (PET), and single-photon emission computed tomography (SPECT) are a few examples providing best-in-class 3D images but require contrast enhancement agent injection. X-Ray and Gamma-Ray modalities may cause radiation hazards. MRI may not be possible if the Patient has implants. MRI, X-Ray and/or Gamma Ray imaging complement each other for soft and hard tissue imaging. Ultrasound sonography has become the most preferred and affordable diagnostic modality but only offers two-dimensional details with a spatial resolution of the order of millimetres [5]. Several other modalities are under development to fill the remaining gaps and reduce usage’s side effects. Electrical impedance tomography (EIT), infrared thermography (IRT), and LASER-based imaging methods are just a few of them [6]–[10]. Electrical impedance tomography provides 2D images with a poor spatial resolution of the order of millimetres [11] but offers a relatively faster temporal resolution. It is used to study respiratory system and breast abnormalities [12]. LASER-based direct imaging modalities are developed for limited ocular imaging applications where at least a translucent biological window is available for internal investigations [13].

X-Ray and Gamma-Ray CT can be used to image the thyroid gland but are not recommended as these diagnostic radiation exposures are associated with high thyroid cancer risk [14] due to the active response of the thyroid gland. Ultrasound provides a low spatial resolution relative to others, and MRI cannot be used for early detection until the Patient is at high risk [15]. Radioactive iodine uptake (RAIU) test is preferred by clinicians as with hyperthyroidism thyroid gland consumes more iodine to produce the T4 hormone. Scintigraphy is suggested for hyperthyroidism and hypothyroidism testing [16], [17]. Any test that involves the use of radioactive tracers has the risk of contamination and can be performed for limited repeatability, especially for pregnant, breastfeeding women, and elderly patients[18], [19].

Besides having discussed modalities, blood tests are clinically preferred to check the severity of the disease. These biochemical-free thyroid tests include tests of T3, T4, TSH, Anti-TPO, and Thyroglobulin levels. Tests for FT3 and FT4 are clinically relevant but have relatively low specific immunoassays [20]. Clinicians recommend having at least one month of an interval between two successive blood tests for hypothyroidism [21].

The Thyroid has a distinct small butterfly shape with two lobes in front of the neck. A prime hormone secreted by the thyroid gland is thyroxin, known as T4, with four iodine atoms. Production of T4 is controlled by the thyroid-stimulating hormone (TSH) secreted by the pituitary gland located at the base of the brain. An imbalance between both hormones may lead to hyperthyroidism/hypothyroidism. Graves’ disease is the most frequently diagnosed disease in India, especially affecting female patients [22]–[29]. It is an autoimmune disease [30] that results in hyperthyroidism or overactive Thyroid. With graves’ disease, the immune system releases thyroid-stimulating immunoglobulin (TSI), an antibody. TSI impersonates thyroid-stimulating hormones (TSH) and stimulates the thyroid gland to produce thyroid hormone in excess. Patients with graves’ disease may acquire malignancies or cancer in the future. The link between Graves’ hyperthyroidism and thyroid cancer is still being explored and studied [31]. Any patient suffering from graves’ disease exhibits primary symptoms such as pain from jaw to ears and mild to moderate fever, etc. These normal symptoms may overlap with other diseases as well. The distinct visual characteristic is swelling of the Thyroid. Swollen eyes are also observed. Afterward, conditions may reach its peak within 3-4 days [32]. It helps to diagnose the symptoms non-invasively; however, any delay requires aggressive treatment protocols. For patients showing early symptoms, RAIU test may be used to image the functioning of the thyroid gland, but it is costly, not entirely harmless, and may not be accessible at every clinic.

Infra-Red thermography provides real-time/direct 2D, non-invasive images with no harmful radiation exposures. The limiting factor is that these images provide sub-surface temperature profiles, but these are shown to have correlations with physiological parameters and can be used for disease diagnostics purposes. At room temperature, human skin emits IR radiation wavelengths ranging from 2-20 µm. IRT (Infra-Red thermography) is used to detect different types of cancers proliferating in the breast [33], liver [34], skin [35], and the brain [36] as the cancerous region found to have relatively higher Temperature and increased blood supply requirements than the other regions. In most cases, thermograms are symmetric in nature [37]–[40]; variation from this symmetry might be a sign of abnormality. It is shown that patients with asymmetric breast thermograms have a higher probability of developing breast cancer within the next five years [41]. Several studies are reported showing the utility of IRT to study Thyroid related diseases. Most of the studies are related to Thyroid Cancer. Few studies have used IRT for Grave disease diagnostics that is also with a very limited cohort size / patient population (not more than 140) [42]–[45].

Besides Hardware, image processing tools are equally important for faster, accurate analysis. Several AI-based image processing algorithms are developed for automatic image segmentation using thermal images [44]-[48].

### Motivation

Thermography is shown to have the potential as a diagnostic tool, especially for Thyroid related diseases. This technique has not been explored much as far as graves’ disease is concerned in Indian demography [49], [50]. A definitive imaging biomarker could not yet be developed due to a lack of clinically validated data. One of the reasons may be the unavailability of (a) affordable and handy thermal cameras along with (b) apt image processing tools at Indian clinics.

An affordable smartphone thermal imager as addon integrated with AI-based image processing soft tool/app is developed. One of the goals is to bring IRT into routine non-invasive check-ups at clinics. As it is already deployed at our clinical partners for testing, the secondary and long-term aim of this article is to provide a simple IRT imaging biomarker for early detection of graves’ disease using the collected data.

To the best of our knowledge, none of the studies reported in the literature includes gender and age-specific categorization of the IRT data despite knowing that the probability of disease distribution is found to be different in female and elderly patients. We have included these two factors in our study to propose an imaging biomarker that is obtained using relatively balanced data.

The next section describes the underlying theory, the methodology followed by the results now.

## Theory

Decision-making parameter, or the “imaging biomarker” to declare if a given patient belongs to the abnormal or healthy category, may depend upon the number of patients used, gender, and age.

M. P. Gopinath and S. Prabu have used the area, variance, and standard deviation of the pixel values under the thyroid region as differentiating parameters [43] [51]. They conclude that area below 50, variance below 5.3, and standard deviation below 35 belong in the category of the healthy person. Tien-Chun Chang and his group showed that the difference in Temperature of the lower eyelid, medial conjunctiva, caruncle, and lateral conjunctiva is significantly higher for the Patient with Grave ophthalmology [52]. Gender and age-specific categorization of the data is not included in any of these studies.

We have used the *difference of average histogram temperature values in control and thyroid region w*.*r*.*t. age and gender* in this work. We have also incorporated the above criteria as different models for comparison purposes.

## Material and methods

Ethical declaration: The study is a collaborative effort between Indian Institute of Technology Roorkee and All India Institute of Medical Science Rishikesh, which is started initially at AIIMSR only. Hardware and software development was completed at IIT Roorkee, and Patient data collection, diagnostics, and clinical validation is carried out at AIIMS Rishikesh, India. This study was approved by the All-India Institute of Medical Science Rishikesh Ethics Committee with reference number AIIMS/IEC/19/997. Informed Consent from patients is taken under Indian Medical Council, Chapter 7, section 14.

A commercially available Thermal imager/camera (a) FLIR E6-XT is used for this study. The developed app must be compatible with other manufacturer models. Images taken by another camera, Testo 882, are also used to include portability. A commercially purchased addon Seek UW-AAA is also used to test the same functionality. This compact model can be attached to any smartphone. We have also assembled our own version of this addon, separately with better control via AI developed app. A window PC-based AI app is also developed on Python and MATLAB platforms. The below sections provide details of Hardware and software / developed applications.

### IR Camera

Following technical specifications are added. These are also available on the manufacturer’s web pages.

All these four IRT cameras also capture images in visual range. These images help to correlate pixel-to-pixel alignment between thermal and visual images if needed.

### Protocols for data collection

After interviewing the patients and reviewing their previous medical history, the thyroid blood tests are suggested. Test reports are reviewed to confirm if the preliminary symptoms are associated with Thyroid. Scintigraphy are performed afterward. These test results categorize the patients into control and hyperthyroid groups. IRT Image is captured such that the temperature profile of the Thyroid (T), chin region (C), and Eye (E) is visible clearly (upright chin-up position). Figure 1 shows one example.

**Fig 1.**
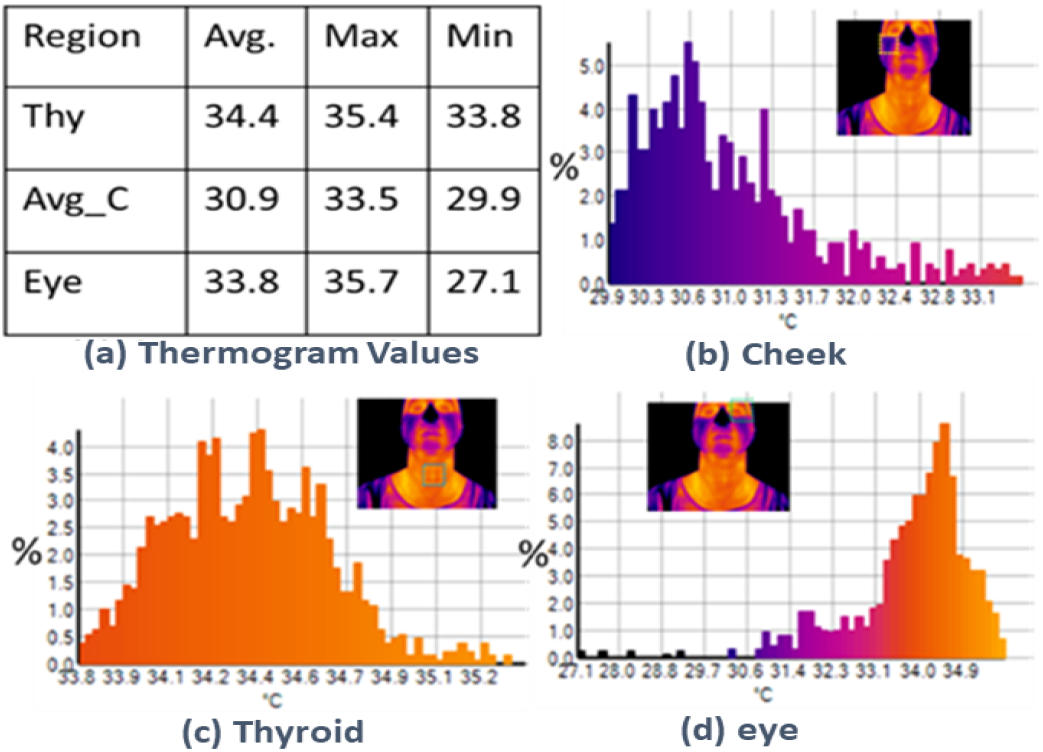
(a) shows quantitative details obtained from histogram inside the rectangles in thermal images, and (b), (c) and (d) show respective histograms (% values vs. Temperature).

Doors and windows of investigation are kept closed to maintain the room temperature between 22 to 26 degrees Celsius. The investigation room did not have any incandescent lighting source. None of the patients wore cosmetics/make-up, any cream or chemicals on the face and neck area. It was inquired that patients did not consume any narcotics and had not gone any physical exercise within the last 30 minutes. It was also ensured that the Patient removed spectacles or goggles before the IRT images session. Ornaments on the neck and ear during the examination are requested to remove. In all cases, we could not ensure if participating /visiting female patients were on the 5th-12th or 21^st^ day of their menstrual cycle.

### Database

In our work, we could only collect 60 Patients, including 44 females (33 with Hypothyroid & 11 healthy as part of the control group) and 16 males (13 with Hypothyroid & 3 healthy as part of the control group).

### Artificial Intelligence Based App

Simple python and MATLAB codes are developed to manually segment the Thyroid, eye, and chin region of the IRT Images and extract the thermogram (histogram from IRT image) values. The manual process is time-consuming and requires certain image processing expertise, inspiring us to opt for an AI-based image processing approach. A typical AI algorithm can be trained to acquire human intelligence to perform a similar task with considerable accuracy.

#### Segmentation Algorithm

Object detection is a computer vision-related exercise. It is designed to detect different features/objects in digital images or movies. Feature detection can be done via already established open-source models, namely R-CNN, Retina-Net, or Single-Shot MultiBox Detector (SSD). These techniques have been found suitable when training data is sparse and/or modelling constraints are present. For feature identification, they require more than one algorithm to run[53]. It significantly increases the execution time, making them unsuitable for app implementation. Alternatively, YOLO (‘You Only Look Once’) is found suitable. The YOLO method uses Convolutional neural networks (CNN) to recognize features in real-time. The technique uses a single forward propagation approach.

#### Model training using YOLO

Darknet framework is used to implement the YOLO algorithm. It is supported by CUDA technology and thus provides computations on GPU that are even faster. A set of 200+ images are used to train the model. Not all the training images belong to the patient data set from the clinic. The size of the used images varies from 240×260 pixels to 640×760 pixels to ensure that any camera model can be used. The throat region is manually selected in all the images, and coordinates are stored in a text file. Both image and text files are used for training the model (Input). The training is then performed on a workstation, HP Z6 128 GB RAM. No differences are found when the same codes are also run-on Google Collab. Around 3252 iterations are performed with an average loss of 0.018530 (As a rule of thumb, once the average reaches below 0.02xxx, the training process is stopped to optimize the processing time). A weight file is obtained after training the model. The throat detection model is then created using this weight file with OpenCV libraries.

#### Segmentation using K-means clustering algorithm

To automatically segment out the unavoidable background features such as edges of clothing in segmented portions and only segment the identified features k-means clustering algorithm is used [54]. This clustering method divides a whole data set into k number of small groups called clusters based on a-likeness. It has two important steps: (a) first is the calculation of the k centroid, and (b) second, a point from every cluster is selected which has the nearest centroid from the respective data point. After it, the distance (Euclidean) of the nearest centroid is calculated. For each cluster new centroid is determined after grouping. Taking the new centroid into account, Euclidean distance is again calculated for the pair of each centre and data point and mapped to the cluster’s points that have minimum Euclidean distance. The member objects and their centroid are unique to each cluster. The centroid of each cluster is the place at which the sum of distances from all of the objects in that cluster is the smallest. Across all clusters, K-means focuses on decreasing the sum of distances between each item and its cluster centroid. The algorithm of the k-means for image size X × Y is as follows:

1. Initialize the k number of clusters and centre of cluster C_k_.
2. Euclidean distance between each pixel P(X, Y) and the centre is calculated as
3. Calculate d for every pixel,
4. Recalculation for the centre using,
5. Repetition of the procedure until convergence,
6. Finally, reshaping of cluster pixels into an image.

Sensitivity analysis is done for values of k. In the first stage, k = 2 is taken to divide each image into two clusters: (a) one for skin and clothing and (b) for the surrounding. This choice is good enough to clearly separate the person from their background. Successively, by setting k =3, the clothing portion is segmented from the person’s exposed part of the skin. This strategy is successfully tested on all data set images from all cameras to discern and segment a temperature difference b/w the skin, clothing, and the surrounding Temperature.

The pixels inside both bounding boxes, i.e., predicted by YOLO and by K-means segmentation, are considered as desired points. The thyroid gland is assumed to be symmetrical in the sagittal plane. After many trials, a suitable region of interest (ROI) for the thyroid gland is found by reducing the size of the bounding box detected by YOLO. This bounding box was reduced by 25% from the left and right, 10% from the top, and 30% from the bottom. All thermal analysis was performed by using points presented in the suitable box (indicated by the black box Fig. 2)

**Fig 2.**
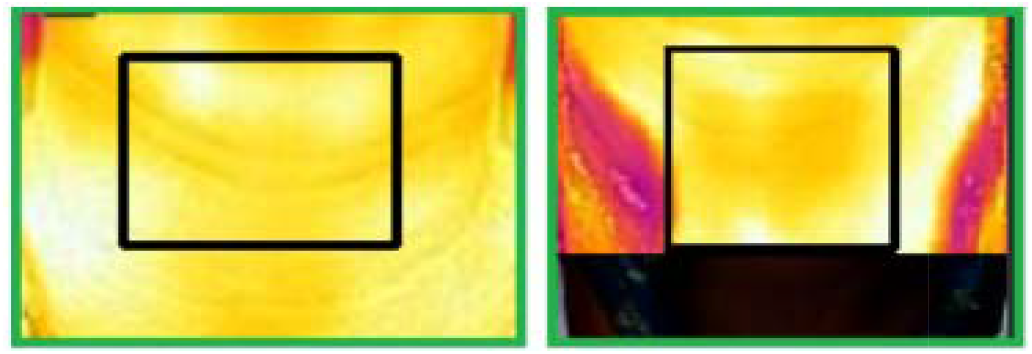
ROI (Throat) predicted by YOLO (Green Box), ROI (Thyroid gland in black box)

### Cheek extraction using face mesh

An open-access library that allows building a multi-model, cross-platform, and applied ML pipeline, namely MediaPipe, is used to create fine face mesh. The desired coordinates are extracted by interpolating the normalized coordinates. It comprises two following steps

1. In the first step, the model scans the entire image and computes face boundaries and locations inside it without additional depth information. A 3D face landmark model that operates on those location points and, using regression, predicts the positions of 3D points and probable surface geometry. The input of this landmark model is a frame of a single RGB camera.
2. In the second step, after the face is correctly cropped, the need for common data augmentations like affine transformations consisting of translations, rotations, and scale adjustments is considerably minimized. It enables the network to devote the majority of its resources to boosting coordinate prediction accuracy. The person’s face is marked with 468 landmark markers (mesh points), which can be used to reach the desired spots with extreme precision. Fig 3 shows the corresponding number of each point in the face mesh. All these images belong to patients and are taken by Thermal Camera.

**Fig 3.**
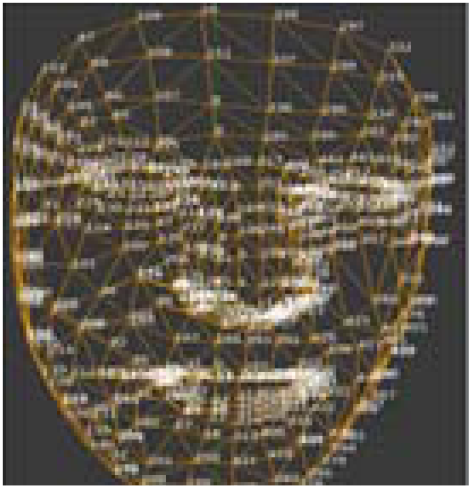
Face Mesh using MediaPipe
3. A rectangular box covering the check area gets selected to extract the cheek pixel values from the corresponding thermal image. The technique gives us control of the shape and size (minimum and maximum value) of this box.

#### Overlapping with thermographs

Please note that the standard IRT camera takes both a thermal image and a standard (RGB) image simultaneously. It is found more suitable to use the regular image than a thermal image to detect face features because (a) a standard (RGB) image has more clarity and well-defined boundaries and (b) better availability of images. As this model is trained on standard (RGB) images, the region of interest from standard (RGB) images to their corresponding thermal images are mapped later. Thermal and visible spectrum images are resized and registered to match their dimension and alignment to overlap during this mapping stage.

#### Temperature Mapping and calibration

Converting the thermal image to a grayscale image for mapping temperatures may result in the wrong analysis as the grayscale values have a many-to-one mapping with RGB values meaning two or more RGB values can give the same grayscale value (Grayscale Value = (R+G+B)/3), which is conflicting as one set of pixel values should point to only one unique Temperature. After extracting the Thyroid and cheek region, Temperature is interpreted from the thermal image. Each thermal image also contains the temperature scale. The app has manual as well as automatic text segmentation options. The RGB pixel values of the thermal image are linearly mapped in between these extracted extremum temperature values. The wavelength associated with the Temperature of each pixel is also noted.

#### Graphical User Interface

All the codes are integrated into a user-friendly and very simple graphical user interface (GUI). One version of this GUI is available as a webpage / fixed URL facilitating an IoT experience. An offline version is also created. To note coordinate and test various buttons for input, upload, and image display, a designer tool provided by pyqt5 is used. It generates a .ui file later converted into a .py file. The working and functions of the widgets are then defined and tested to check for errors. Using pyinstaller, this .py file is finally converted into an executable (.exe) file. A supplementary readme file is provided to operate the app.

## Result and Discussion

We have analysed segmented thermal data from sixty patients of different age groups and gender; among them, thirty-three females and thirteen males are found to have hyperthyroid, confirmed by standard Blood and scintigraphy tests. A case study data is presented in Fig. 4. It is observed that for a healthy / control person, the difference between imaging marking values of the thyroid region (T1) and average values of both cheek regions (C1 and C2) is low; however, for a patient with hyperthyroidism, it was high (Diff_C). Both people belong to the same age group. Using the same marking values, the difference between region T1 and C2 (Diff_G) is high; however, for healthy people, it is less.

**Figure 4.**
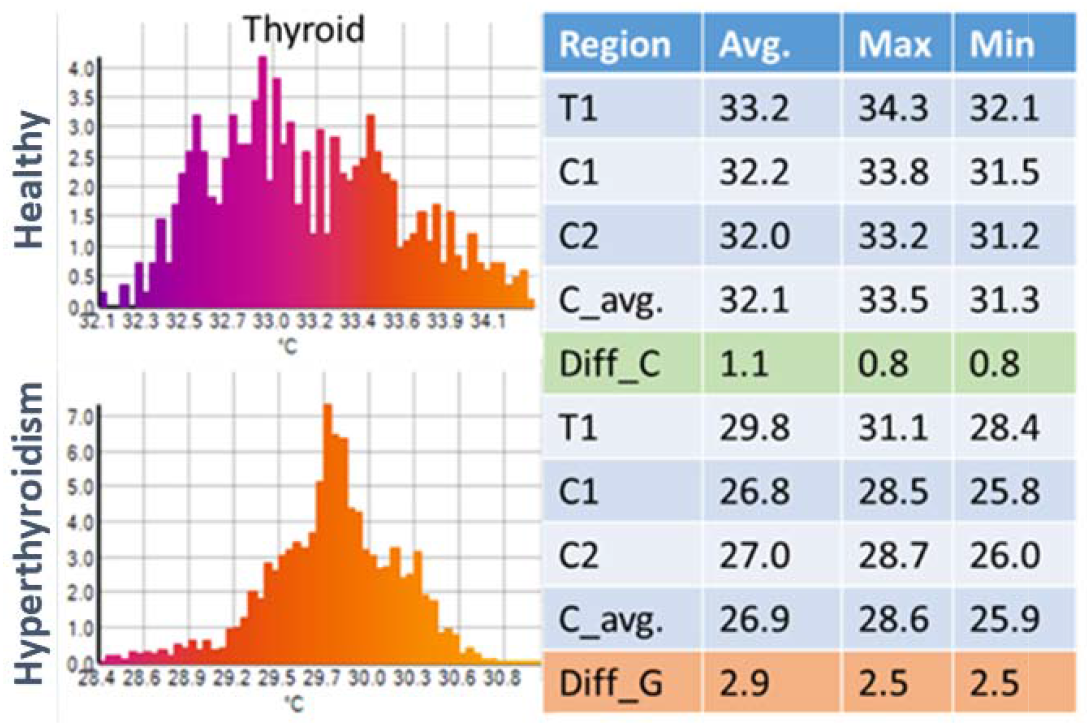
Quantitative analysis of a healthy person and Patient

Difference values (maximum, minimum, average and mean) between thyroid and control region v/s age are plotted for all female visitors now in Figs. 5(a) and 5(b), respectively. Male data analysis is skipped for brevity purposes; however, it shows a similar outcome with relatively moderate correlations. No smoothening is performed in data analysis. The Minimum and Maximum values obtained from thermal images are the weakest mathematical parameter as they do not show any significant difference. The functional analysis in Fig. 11 presents a stark difference between the thyroid group and the control group w.r.t age.

**Figure 5.**
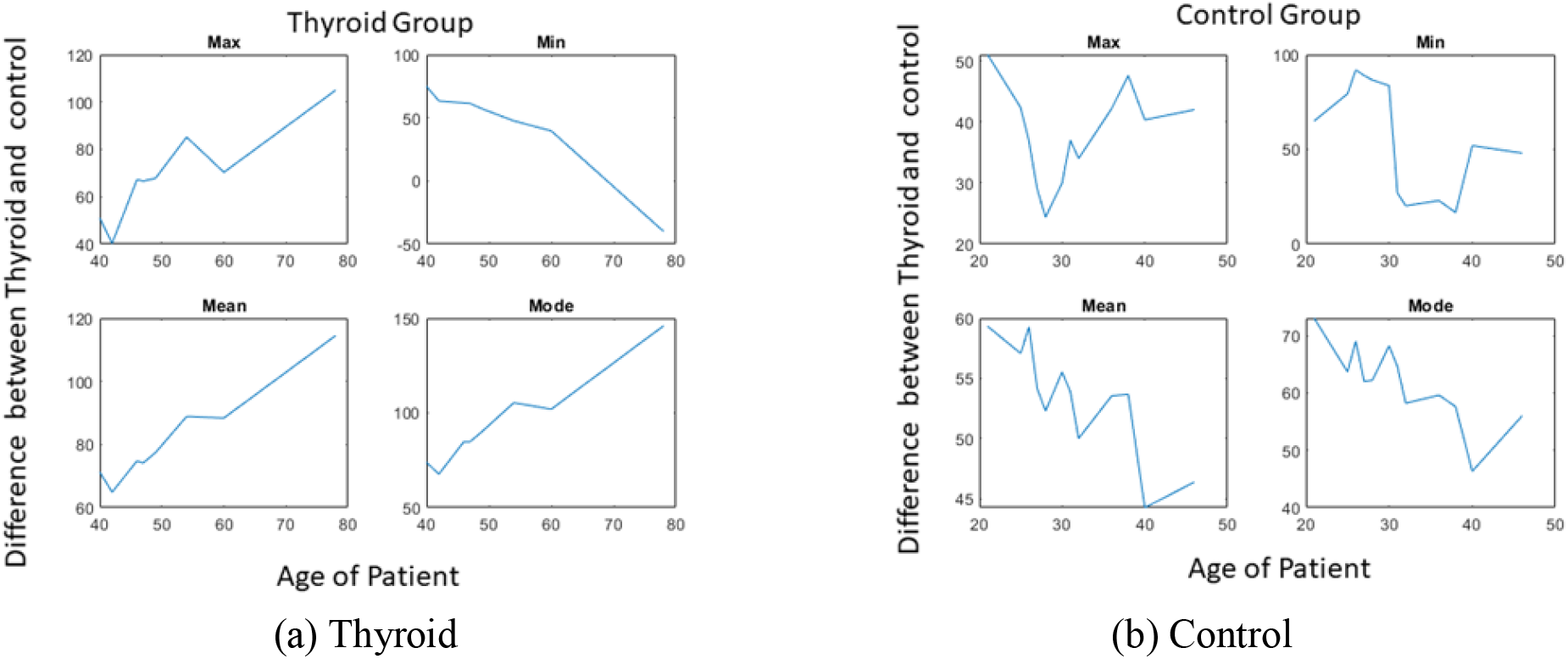
Thermal Image Analysis for Female Volunteers

The mean value of the temperature profile between the thyroid region and cheek region obtained from a graves’ disease female patient has a positive gradient w.r.t. age group. The same gradient is negative in the case of control or healthy volunteers. Figure 6 shows one example when a new patient data / thermal image is tested using this analysis. The normalized mean values and their linear fit from control and graves’ diseases is plotted in the same graph. The difference mean value between thyroid and cheek region from one new patient data (not used either in control or graves plots) is also overlapped. The euclidean distances of this new point from the linear fits from the control group and graves’ group, C1 and T1 are estimated. It assumed that if the point is near the control group (C1< T1), the person whose image is fed to the analysis is not suffering from graves’ disease. However, if the point is near to the linear fit of the graves’ disease plot (T1<C1), then the volunteer is considered a patient. The severity is decided according to the nearness. In case T1 is equal to C1, the case is declared as borderline. In this analysis, the fit can also be non-linear based upon the minimum goodness of fit.

**Fig 6.**
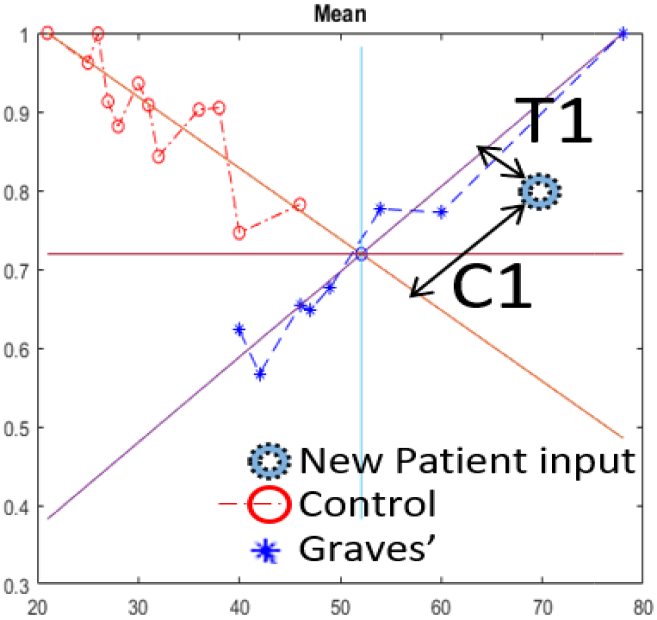
Thermal Imaging Biomarker

The outcome of this analysis depends upon the richness of the statistical data fed to it. An app is developed, integrated with in-house developed IR smartphone addon/hardware, and deployed at the clinical partner’s end to enrich. The screenshot of this app is shown in Fig. 7. It has an in-built option to upload both images (IR and visible range) along with entry options about patient details. It is a process to generate the location of fresh data (shown in the Red star marker), providing its assessment to the clinician. Clinicians can correct or verify if the diagnosis is correct (if other test data is available). This correction can be integrated into the database to update the AI weights for forthcoming diagnoses. The dedicated option of “diagnostics input by clinician” is given. Another commercially available smartphone addon, a compact and relatively cheaper IR Camera (sensor technology model HTPA120×84d), is tested instead of a complicated and bulky standard thermal camera to carry out the same analysis. It produces the same analysis however drains the battery of the mobile unexpectedly faster. It also has its own dedicated app. We have developed an in-house addon similar to this. It requires a user to use two apps; (a) to control the thermal addon and take thermal images and visible images. Afterward, the user has to upload these images into the AI IRT diagnostics app for analysis. We have fabricated our own thermal smartphone addon similar to the one that we purchased commercially. We have used MLX90640 technology interpolated to high resolution. The technology (without smartphone integration) is reported earlier. This in-house developed addon has provided us better control to acquire a thermal and visual image in a single push of a button by immediately processing and providing the result. It is relatively a cheaper option. The single app control improves the user experience and the speed of analysis and reduces any manual error to uploading the wrong set of images. The details are given in Table 1.

**Fig. 7.**
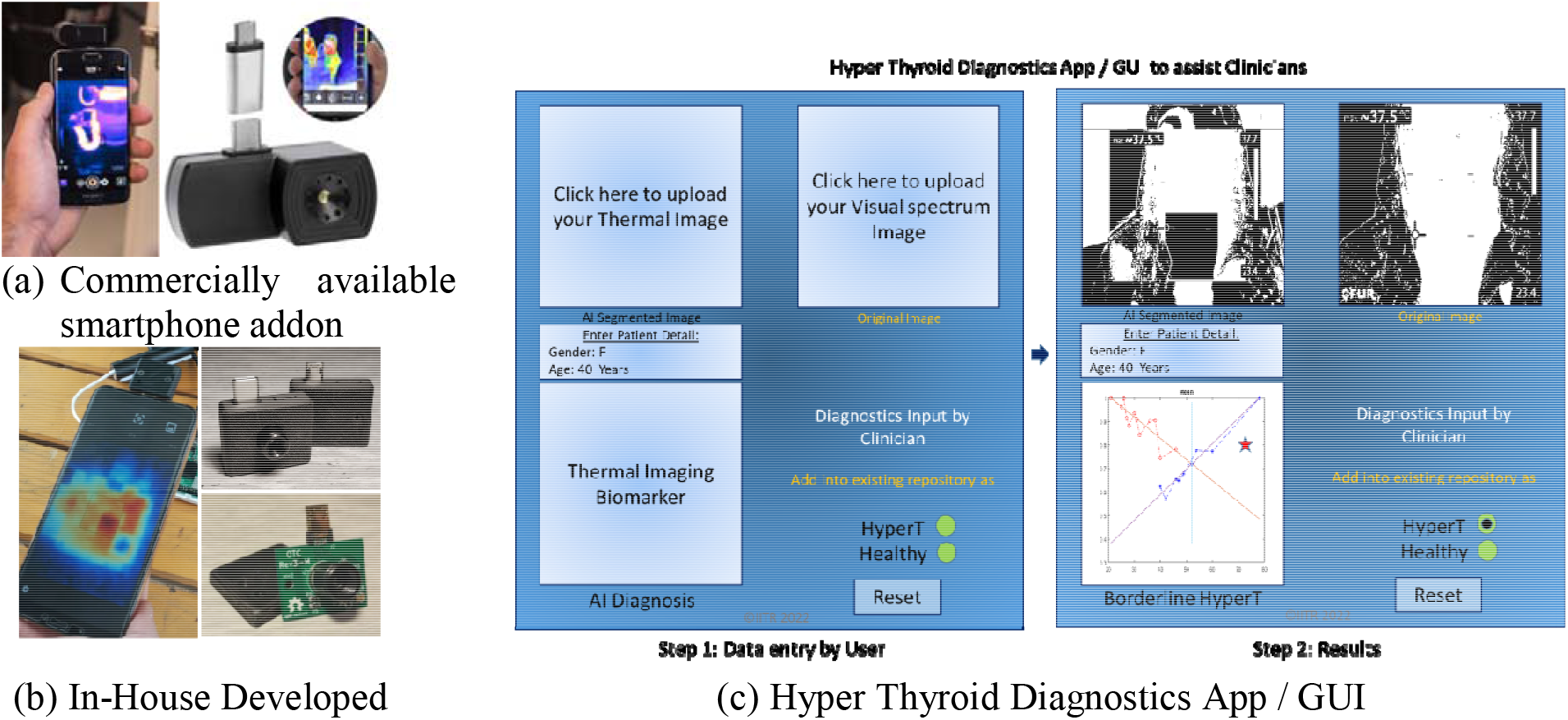
Point-of-Care Thyroid Pre-Diagnostics Tool to assist Clinicians.

**Table 1:** Details of IR Camera used in this study

| S.No. | Tech. Spec. | FLIR E6-XT | Testo 882 | Seek | Add-on |
| --- | --- | --- | --- | --- | --- |
| 1. | Camera size (mm) | 244 × 95 × 140 | 152 x 108 x 262 | 25.4 x 44.45 x 20.32 |  |
| 2. | Form factor | Handheld Camera | Handheld Camera | Smartphone add-on / plugin | Smartphone add-on / plugin |
| 3. | IR Resolution | 240 × 180 pixels | 320 x 240 pixels | 206 x 156 pixels |  |
| 4. | Thermal Sensitivity | <60 mK | <50 mK at +30°C | NA |  |
| 5. | Type of detector | Uncooled microbolometer | NA | Microbolometer |  |
| 6. | Image frequency | 9 Hz | 33 Hz | <9 Hz |  |
| 7. | Spectral range | 7.5–13 μm | 7.5 to 14 μm | 7.5 - 14 μm |  |
| 8. | Accuracy (10°C to 35°C) | ±2°C, | ±2°C, | NA |  |

## Discussion

Cost-effectiveness can help to provide this point-of-care device to a patient for self-assessment before even starting to fix an appointment at the clinic. This study has two limitations: firstly, due to cost optimization, the point-of-care device has VGA level resolution. It may lead to erroneous segmentation of butterfly shape thyroid. Thyroid and control region shape is kept square in this study. However, a high-resolution camera is also used to verify the outcome. Second, even if the segmentation of the Thyroid is accurate and verified (with gold standard tests) patient population is sparse.

## Conclusion

We have developed IR thermography-based point-of-care device to pre-test the Thyroid related clinical conditions. The device has an AI-based smartphone app along with a hardware/addon. The app utilizes a simple and novel difference-based imaging biomarker. Qualitatively as well as quantitatively, there appears to be a difference in IR images of patients with known hyperthyroidism. The results are supported by gold standard scintigraphy and blood tests. Here are pointwise conclusions:

1. AI is used only to segment the thyroid and control region. It is not used to develop imaging biomarkers for diagnostics as patient data is sparse right now and may become biased w.r.t gender or age after prolonged use.
2. The sparseness, however, is expected to be reduced as the point-of-care device may help gather more data in the near future.
3. The age of the Patient is an affecting factor, and more samples per Age group will increase the reliability of the outcome. The difference (Max value) between the Thyroid and control region remains low (<50) for patients in the control group as compared to patients suffering from hyperthyroid (>50). This difference, however, seems to be significantly larger if the Patient is old.
4. Control and hyperthyroid have different behaviour and lie in different quadrants, which can help clinicians to conclude whether the Patient is healthy or not. If results suggest that a person is suffering from the Grave disease clinician can suggest the standard tests; otherwise, can ask to revisit after a few weeks or so, saving expense on the patient side. More data will make this study more reliable.

These results are encouraging to further study a larger number of patients for statistically sound conclusive.

## Data Availability

All data produced in the present study are available upon reasonable request to the authors.

## Acknowledgments

Vaishali Sharma is thankful to the CSIR Government of India for fellowship, MG is thankful to Mr. S. Kandari and Mr. S. Prakash for app development and to Dr. S. Shakya assisting in early-stage data processing analysis.

## CRediT authorship contribution statement

VS: Investigation and Writing; MG: Methodology, Supervision, Investigation, Software, Writing, Visualization; SS: Investigation, Software, AK: Investigation, VKD: Data Acquisition, and Clinical Diagnostics and Supervision.

## Supplementary File

User Guide steps for GUI are as follows:

1. Users need to upload both regular and thermal images in the first step,
2. In the second step, users need to provide important information about the Patient, like age, gender, and minimum and maximum Temperature of the regular and thermal image.
3. The model will run, and graphs will be generated by clicking the RUN button in the next step. A red dot will show new data.
4. In the final step, the user will have an option to save the study for a healthy or non-healthy person.

